# Differential causative effects of germline pathogenic variants in *MUTYH* and *PALB2* in a patient with colorectal polyposis and breast cancer

**DOI:** 10.64898/2026.05.15.26352890

**Authors:** José Camacho-Valenzuela, Dylan Pelletier, Paz Polak, Lili Fu, Nancy Hamel, Céline Domecq, Afrida Ahmed, Carla Daniela Robles-Espinoza, William D. Foulkes

## Abstract

**Purpose:** Patients carrying Germline Pathogenic Variants (GPVs) in multiple cancer susceptibility genes (CSGs) can be described within the context of Multi-locus Inherited Neoplasia Allele Syndrome (MINAS). The role of each GPV is typically interpreted based on clinical phenotypes. Here, we used tumor sequencing, particularly mutational signatures, to investigate the contribution of GPVs in *MUTYH* and *PALB2* to colorectal polyposis and breast cancer in a single patient at a molecular level.

**Methods:** We analyzed tumor sequencing data, including mutational signatures and genomic scars, of a breast tumor and a colorectal polyp from a patient with biallelic GPVs in *MUTYH* and a heterozygous GPV in *PALB2*.

**Results:** The colorectal polyp showed a dominant contribution of *MUTYH*-associated Base Excision Repair deficiency (BERd) mutational signatures, with no evidence of Homologous Recombination Repair Deficiency (HRD). In contrast, the breast tumor showed both *MUTYH*-driven BERd and HRD-associated signatures, including SBS3, ID6 and an elevated HRD score, despite the absence of a detectable second hit in *PALB2*. These findings suggest a differential contribution from the CSGs, with *MUTYH* contributing to both lesions and *PALB2* contributing specifically to the breast tumor. The observed pattern does not align with the additive or synergistic models described in MINAS.

**Conclusions:** Our study provides evidence that mutational signatures can elucidate the contribution of multiple CSGs to tumorigenesis within a single patient. These findings extend current interpretations of MINAS beyond additive or synergistic phenotypes, which may help to better understand tumor etiology, with potential clinical implications, including eligibility for targeted therapies.

## Introduction

Colorectal polyposis is defined by the presence of >10 cumulative adenomatous polyps in a lifetime^1^. Numerous underlying genetic syndromes have been identified as causing adenomatous polyposis, with genetic testing being warranted based on specific criteria including family history, presence of 20 or more adenomas, and specific findings on pathology^1-3^.

Adenomatous polyposis may be caused by various genetic syndromes, including classic or attenuated Familial Adenomatous Polyposis (FAP) caused by heterozygous germline pathogenic variants (GPVs) in *APC*^4^. Classic FAP, an autosomal dominant condition, is usually marked by the presence of more than 100 colorectal adenomas at a young age, while attenuated FAP features less than 100 colorectal adenomas^5^. In addition, polymerase proofreading-associated polyposis, caused by heterozygous GPVs in *POLE* or *POLD1* can lead to a polyp burden similar to attenuated FAP^6^.

Among autosomal recessive conditions, biallelic GPVs in *MUTYH* cause colorectal polyposis, typically with about 10 to 100 polyps and onset ranging from the 20s-70s^7^. Less frequently, biallelic GPVs in *NTHL1, MSH3*, or *MBD4* can also cause gastrointestinal adenomatous polyposis, with a similar polyp burden^8-10^. Constitutional Mismatch Repair Deficiency (CMMRD) syndrome may lead to rather adolescent-onset polyposis^11^.

In hereditary breast cancer, genetic predisposition is often linked to specific tumor subtypes. Triple negative breast cancer is mostly associated with GPVs in *BRCA1/2, PALB2*, or *RAD51C*/*D*^12,13^. Estrogen receptor-positive breast cancer is associated with GPVs in *ATM, CHEK2, PALB2, or BRCA2*^12,13^. HER2-positive breast cancer is frequently associated with *TP53* while lobular breast cancer is linked to *CDH1*^14,15^.

Only a few associations have been made between predisposition to breast cancer and polyposis. For example, in addition to polyposis, GPVs in *NTHL1* predispose to a wide variety of malignant lesions including pancreatic, brain, hematological, as well as breast cancer, with a median age of onset of approximately 48 years old^16-18^.

In contrast, the association between monoallelic or biallelic GPVs in *MUTYH* and breast cancer risk remains unclear, with inconsistent findings across studies^19,20^. Beyond polyposis and colorectal cancer, biallelic *MUTYH* GPVs are known to predispose to duodenal, ovarian, and bladder cancers^18^. Base Excision Repair deficiency (BERd)-associated mutational signatures have been reported in a pancreatic ductal adenocarcinoma from a patient with biallelic GPVs in *MUTYH*^21^. Similarly, breast cancers from two individuals with monoallelic *MUTYH* GPVs and somatic loss of the second allele showed evidence of BERd, including modest presence of COSMIC SBS18/SBS36, and very limited contribution of signatures associated with Homologous Recombination Deficiency (HRD)^21^. However, these tumors also showed substantial age-related or APOBEC-associated signatures, making the role of *MUTYH* in tumor development uncertain^21^.

With the increasing number of individuals receiving germline genetic testing, co-occurrence of GPVs in multiple Cancer Susceptibility genes (CSGs) is being identified more frequently^22,23^. This phenomenon has been termed Multi-locus Inherited Neoplasia Allele Syndrome (MINAS), referring to individuals carrying GPVs in two or more CSGs^22^. This phenomenon had been reported at least as far back as 1996^24^, but was not specifically named as MINAS. In most cases, each CSG contributes independently to its associated tumor phenotype (referred to as additive model)^23,25^. However, some studies have reported individuals with MINAS developing tumors not typically associated with each individual CSG (described as synergistic model)^23^. Clinically, multiple primary tumors are frequently present in individuals with MINAS, likely due to the presence of at least two germline alterations^23^. Nevertheless, the biological mechanisms involved in MINAS and the contribution of each GPV to tumor development remain under-investigated.

In this context, we report a patient with *MUTYH*-associated polyposis (MAP) caused by biallelic GPVs in *MUTYH* who developed breast cancer in her 40’s and was subsequently found to carry a GPV in *PALB2*, another well-known CSG. We investigated whether the breast cancer and polyposis were driven by *PALB2*-related HRD, *MUTYH*-related BERd, or neither and how this case fits within the additive versus synergistic models in MINAS. To address this question, we analyzed tumor sequencing data, including mutational signatures, from the breast tumor and a colorectal polyp.

## Materials and Methods

### Patient samples and whole-exome sequencing

Peripheral blood and formalin-fixed paraffin-embedded tumor blocks were collected for analysis. Genomic DNA extraction and whole-exome sequencing (WES) were performed as previously described^26^. Variant filtering and prioritization followed an approach adapted from a previous study^26^. Tumor mutational burden (TMB) was defined as the number of somatic non-synonymous coding mutations per megabase of coding sequence^27^ (see **Supplementary Information**).

### Mutational signature, genomic scar and copy-number analysis

Single base substitution (SBS) and INDEL mutational signatures were analyzed using MutationalPatterns^28^ and SigProfilerAssignment (v0.1.9)^29^. Somatic INDELs were manually reviewed in the Integrative Genomics Viewer to exclude potential artefacts. HRD-associated signatures SBS3 and SBS8 were assessed using SigMA^30^. Genomic scar analysis and copy-number profiles were analyzed as previously described^26^ (details in **Supplementary Information**).

## Results

### Case presentation

The patient, of North African ancestry, was referred to gastroenterology in her 30’s due to a family history of colorectal cancer in one brother in his 30s and duodenal cancer in another sibling in their 40s (**Table 1** and **Supplementary Figure 1**). The same year, colonoscopy identified 15 polyps. Pathology showed multiple tubular adenomas and one tubulovillous adenoma. Mismatch repair protein immunohistochemistry showed retained nuclear expression of all four proteins (MLH1, MSH2, MSH6 and PMS2). She subsequently underwent germline testing with a polyposis and colorectal cancer gene panel (**Supplementary Table 1A**), which identified the *MUTYH* GPV (NM_001128425.1) c.1227_1228dupGG; p.Glu410Glyfs43 in the homozygous state, with a ClinVar classification of pathogenic (Variation ID: <u>127831</u>). Follow-up colonoscopies consistently identified approximately 10–20 polyps, while routine upper endoscopy showed no polyps.

Five years later, she noticed a mass on her left breast. On exam, she was noted to have left lymphadenopathy. Subsequent biopsy revealed ER/PR-positive, HER2-negative infiltrating ductal carcinoma and metastatic carcinoma in the left axillary lymph node. She underwent chemotherapy consisting of paclitaxel, adriamycin and cyclophosphamide. Given her young age, she was referred for hereditary breast and ovarian cancer genetic testing (**Supplementary Table 1B**), which identified a germline heterozygous exon 5 deletion in *PALB2* (NM_024675.3), which was confirmed clinically by Multiplex Ligation-dependent Probe Amplification (**Supplementary Figure 2**). No GPVs were observed in the well-established breast and ovarian CSGs *BRCA1/2*. This guided the decision for the patient to undergo a bilateral mastectomy following neoadjuvant treatment. Following surgery, she was staged as pT1c pN1a M0 and received adjuvant radiotherapy and abemaciclib. The patient is currently in remission and on hormone therapy, four years following her surgery.

**Table 1.** Clinical and germline information of the patient. *Key: The ages are approximate to protect patient’s identity. Abbreviations: MAP (*MUTYH*-associated polyposis); MMR (Mismatch repair); IHC: Immunohistochemistry; HBOC (Hereditary breast and ovarian cancer).

|  |  |
| --- | --- |
| **Ancestry** | North African |
| **Sex** | Female |
| **Family history** | Sibling with colorectal cancer at 26-30 years*<br>Another sibling with duodenal cancer at 41-45 years*<br>No known consanguinity |
| **Age at diagnosis (multiple polyps)** | 31-35 years* |
| **Age at diagnosis (breast cancer)** | 41-45 years* |
| **Polyp:**<br>**Pathology and MMR IHC** | Tubular adenomas and one tubulovillous adenoma<br>Normal nuclear expression of MLH1, MSH2, MSH6 and PMS2 |
| **Breast tumor:**<br>**Histology and receptor status** | Infiltrating ductal carcinoma<br>ER-positive, PR-positive, HER2-negative |
| **Germline clinical testing**<br>**(polyposis gene panel)** | *MUTYH* (NM_001128425.1):<br>c.1227_1228dupGG; p.Glu410Glyfs*43; homozygous |
| **Germline clinical testing**<br>**(HBOC gene panel)** | *PALB2* (NM_024675.3), exon 5 deletion,<br>confirmed by MLPA; heterozygous |

### Molecular investigations

Germline WES confirmed the homozygous GPV in *MUTYH* (c.1227_1228dupGG), with a VAF of 96%, consistent with biallelic inactivation. No somatic exonic variants were observed in *PALB2* in neither tumor nor in *BRCA1/2*, and copy-number analysis showed no evidence of LOH at the *PALB2* locus in the polyp or the breast tumor (**Supplementary Figure 3**). Somatic WES in the polyp identified potential disease-causing somatic variants in cancer-associated genes, including a nonsense mutation in *APC* (p.E425*, VAF 5.7%) and a missense mutation in *KRAS* (p.G12C, VAF 8.6%), both of them confirmed via Sanger sequencing. In the breast tumor, we identified a somatic nonsense variant in *GATA2* (p.E224*, VAF 10.5%), among others (**Supplementary Table 2A**). The overall somatic mutation burden was low in both samples. Specifically, a total of 179 and 116 exonic and splice-site somatic variants were identified in the polyp and breast tumor, respectively. Based on the subset of non-synonymous coding variants, TMB was estimated at 3.9 and 2.4 somatic mutations per megabase of coding sequence in the polyp and breast tumor, respectively, consistent with low mutational load in both samples.

To further evaluate the presence of HRD, genomic scar analysis revealed an HRD score of 18 in the colorectal polyp, well below the reported thresholds of ≥42 and ≥33 (see **Supplementary Information**), indicating no evidence of HRD. In contrast, the breast tumor exhibited an HRD score of 33 which, indicates an appreciable level of genomic scars consistent with HRD.

Mutational signatures provided additional insights into the underlying mutational processes. Dominant BERd signatures were observed in the polyp (**Figure 1A** and **Supplementary Table 2B**), whereas the breast tumor exhibited notable contributions from both BERd and HRD-associated signatures (**Figure 1B** and **Supplementary Table 2B**). Specifically, in the colorectal polyp, SigMA and SigProfilerAssignment showed a dominant contribution (about 70%) of the *MUTYH*-associated SBS18, while MutationalPatterns identified SBS36 (also associated with *MUTYH* biallelic inactivation) as the unique contributing SBS signature. No evidence of HRD-related SBS or INDEL signatures was observed. The reconstructed SBS profile showed high cosine similarity scores (MutationalPatterns: 0.96; SigProfilerAssignment: 0.97), and a slightly lower cosine similarity value for the reconstructed ID profile (MutationalPatterns: 0.88, no ID signature observed with SigProfilerAssignment), as shown in **Supplementary Table 2C**.

In the breast tumor, SigMA classified this sample as SBS3 positive with high confidence (mva score of 0.95, additional SBS3 metrics in **Supplementary Table 2D**), with a moderate contribution of this HRD-associated signature (about 20%), alongside a dominant contribution of the *MUTYH-*associated SBS18 (approximately 50%). Minor contributions of the APOBEC signatures SBS2 and SBS13 were also observed. SigProfilerAssignment identified a dominant contribution of the *MUTYH*-associated SBS36 (about 60%), alongside a moderate presence of APOBEC signatures (approximately 20%). Both SigProfilerAssignment and MutationalPatterns identified the HRD-related INDEL signature ID6, which was the unique INDEL signature detected by SigProfilerAssignment, and a major contributor in MutationalPatterns (about 90%). In addition, MutationalPatterns identified a remarkable contribution of SBS39 (approximately 40%). The reconstructed SBS profile showed high cosine similarity scores (MutationalPatterns: 0.92; SigProfilerAssignment: 0.93), and more moderate cosine similarity values for the reconstructed ID profile (MutationalPatterns: 0.81; SigProfilerAssignment: 0.68).

**Figure 1:**
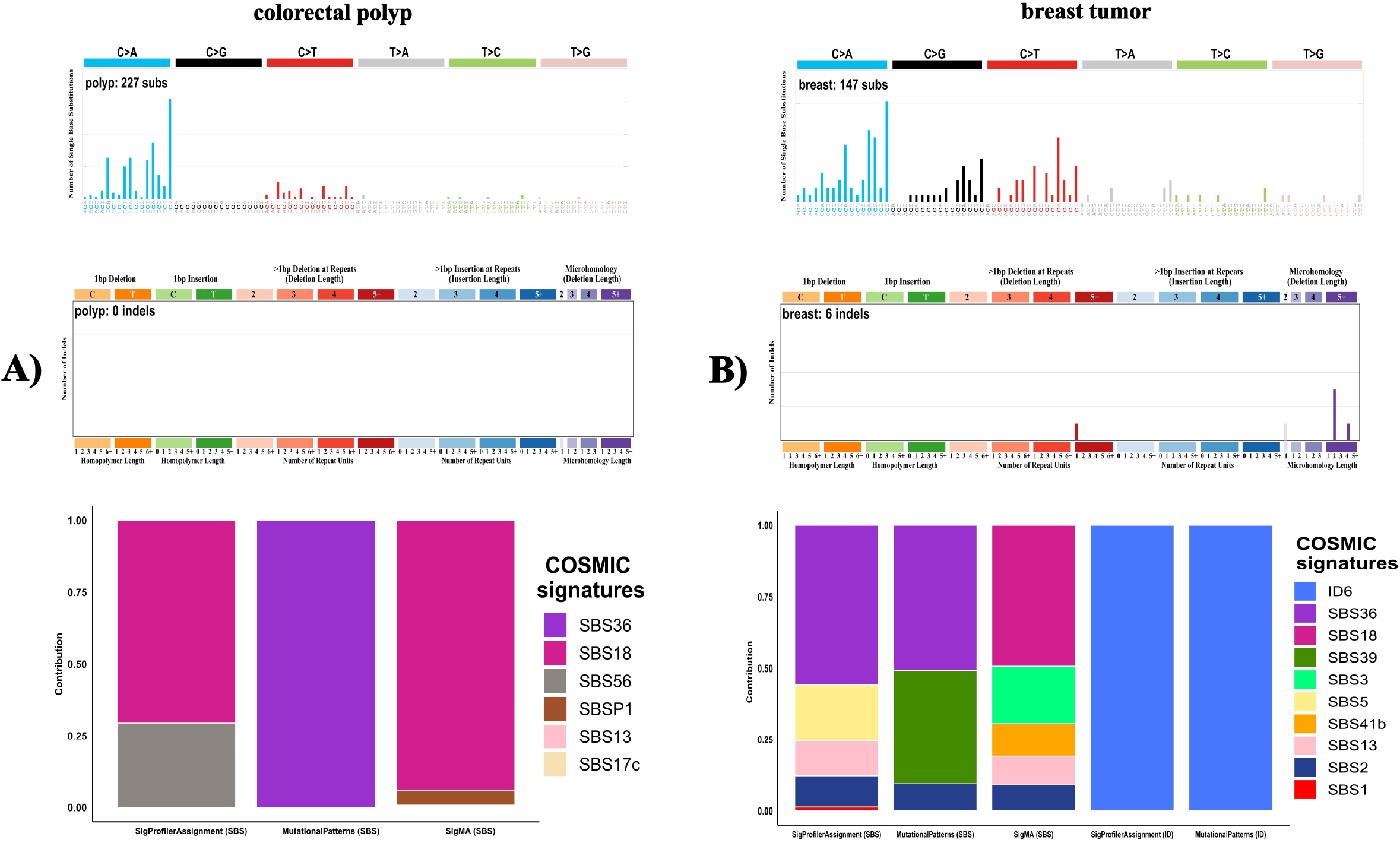

## Discussion

In this study, we present the case of a patient carrying two co-occurring GPVs in the well-established CSGs *MUTYH* and *PALB2*, who developed colorectal polyposis consistent with the expected *MUTYH*-associated phenotype, but with evidence of a differential contribution of the two genes in the breast tumor, as supported by mutational signature analyses. Although no somatic second hit in *PALB2* was detected by WES, the collective presence of HRD-associated mutational signatures and genomic scars in the breast tumor supports an HRD phenotype suggestive of an underlying biallelic inactivation of this gene. Hence, the absence of a detectable second hit in *PALB2* likely reflects the limited sensitivity of WES to capture alterations potentially acting as second hits, including deep intronic variants^31^, promoter methylation events^32^, or structural variants, as well as reduced sensitivity in low tumor purity (**Supplementary Figure 3**). Although the reconstructed ID profiles showed more moderate cosine similarity scores in the breast tumor (as described in Results), these values should be interpreted in the context of the observed low tumor purity, limited number of INDELs and the intrinsic limitations of analyzing mutational signatures with WES, which may affect the accuracy of cosine similarity scores. Nevertheless, the identification of ID6 together with SBS3 further supports the HRD phenotype.

It has been proposed that phenotypic consequences of MINAS may be either additive, where each CSG contributes independently to its expected phenotype, or synergistic, where the combination of CSGs may lead to more severe phenotypes like earlier disease onset or tumors not typically associated with the observed genotype^23^. The pattern observed in this patient fits neither model fully. The polyp etiology is clear and consistent with *MUTYH* biallelic inactivation, supported by dominant contributions of BERd-related SBS18 and SBS36 and the absence of HRD signatures. However, the breast tumor appears to be explained by a combined contribution of BERd and HRD, as supported by *MUTYH*-associated SBS18/SBS36, and HRD signatures consistent with *PALB2*-deficiency, including SBS3, ID6, SBS39 (the latter of which has also been recently associated with HRD, including in breast tumors with germline and somatic mutations in *BRCA1/2* and *PALB2*^33^), and an appreciable genomic scar-based HRD score. These patterns suggest a differential contribution that differs from the additive and synergistic models described in MINAS^23^.

A prior study reported two breast cancers in patients with monoallelic *MUTYH* GPVs with somatic loss of the wild type allele, consistent with biallelic inactivation^21^. In both breast tumors, SBS18/SBS36 were present but with modest contributions. In the first patient, the clock-like SBS1 was dominant and no alternative genetic driver was identified. In the second case, a co-occurring GPV in *CHEK2* (a well-established breast CSG) with somatic loss of the wild type allele was observed, which may represent a more likely explanation for breast tumor development. In overall, the authors concluded that, although the role of *MUTYH* in breast tumorigenesis remains uncertain, *MUTYH*-associated events may have contributed to tumor evolution^21^. In contrast, our case exhibited a much clearer presence of *MUTYH*-driven BERd and *PALB2*-associated HRD phenotypes, consistent with differential contributions of these CSGs.

Another study reported the somatic *KRAS* transversion c.34G>T observed in our case in seven MAP patients with biallelic *MUTYH* GPVs^34^. Given that transversions are a characteristic feature of *MUTYH* biallelic inactivation, the presence of this same somatic mutation in the colorectal polyp of our MAP case, alongside dominant *MUTYH*-associated mutational signatures, supports a key role of *MUTYH*-driven BERd in this lesion.

In the context of our findings, *MUTYH* deficiency alone appears sufficient to drive polyposis, but unlikely to drive breast tumorigenesis independently. The observed co-occurrence of BERd and HRD mutational signatures instead suggest that, in the context of *PALB2* deficiency, *MUTYH*-driven BERd may contribute to breast cancer. These findings have clinical significance. The presence of HRD-associated genomic features in the breast tumor points to potential eligibility for treatment with Poly ADP-ribose polymerase (PARP) inhibitors. Additionally, the identification of GPVs in *MUTYH* and *PALB2* has clinical implications for cascade testing and cancer risk assessment for the patient’s relatives.

In summary, to the best of our knowledge, this is the first report of a patient with polyposis and breast cancer carrying GPVs in *MUTYH* and *PALB2*, in whom mutational signatures revealed a differential contribution to tumor development. While validation in larger cohorts is required, these findings may extend current interpretation of MINAS beyond additive or synergistic models and highlights the value of mutational signatures to investigate the causative effects of GPVs in multiple CSGs within a single patient.

## Data Availability

De-identified relevant data supporting the findings of this study are included in the manuscript and supplements. Raw sequencing data are available upon reasonable request from the corresponding author.

## Acknowledgments

We thank Zhen Shen for her valuable support with the collection of clinical data.

## Authors contributions

Conceptualization: W.D.F, N.H., C.D.R.E.; Data curation: J.C.V.; Formal analysis: J.C.V.; Funding acquisition: W.D.F., N.H.; Investigation: W.D.F., D.P., L.F. AA., CD.; Project administration: W.D.F., N.H.; Resources: L.F., AA., CD.; Supervision: W.D.F., N.H., C.D.R.E, P.P.; Visualization: D.P.; Writing original draft: J.C.V., D.P.

## Ethics declaration

Written informed consent was obtained from the patient. This study was approved by the McGill University Health Centre Research Ethics Board. Project No: MP-37-2019-4865.

