## Supplementary figures and images for "Differential causative effects of germline pathogenic variants in *MUTYH* and *PALB2* in a patient with colorectal polyposis and breast cancer"

### supplementary figure 1

**Supplementary Figure 1**

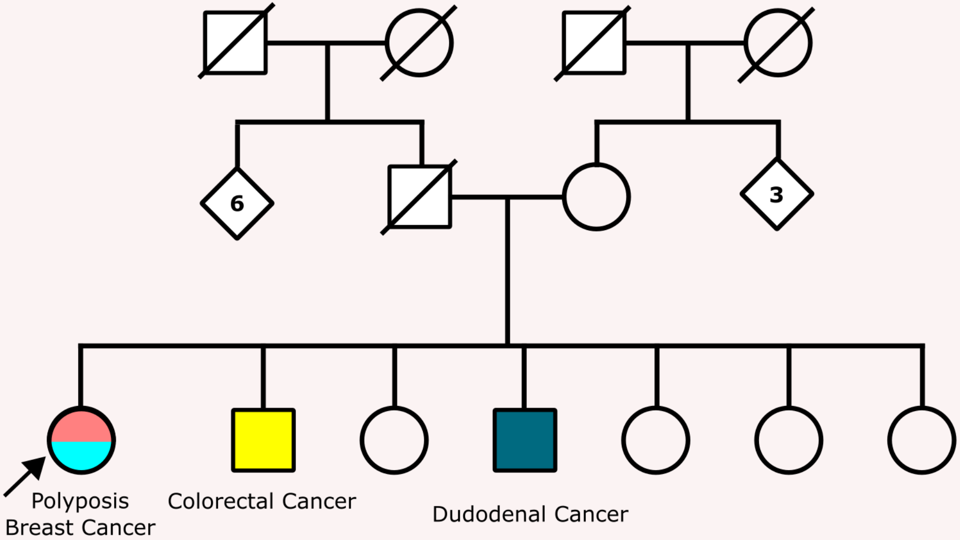

### supplementary figure 3

#### Supplementary Figure 3

##### A) Colorectal polyp (cellularity: 0.1)

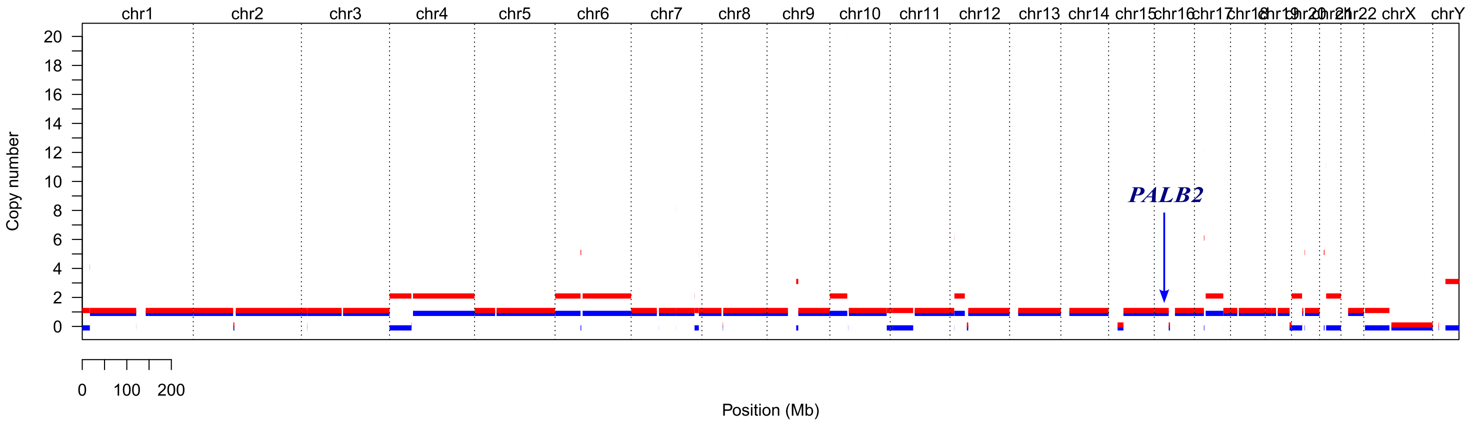


##### B) Breast tumor (cellularity: 0.2)

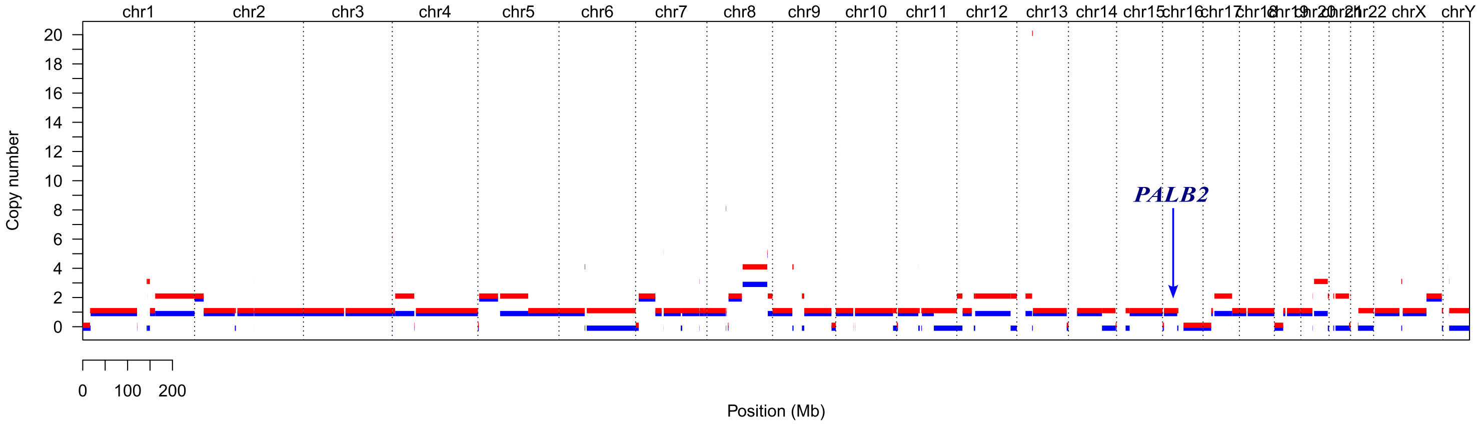
