## supplementary figure 2 for "Differential causative effects of germline pathogenic variants in *MUTYH* and *PALB2* in a patient with colorectal polyposis and breast cancer"

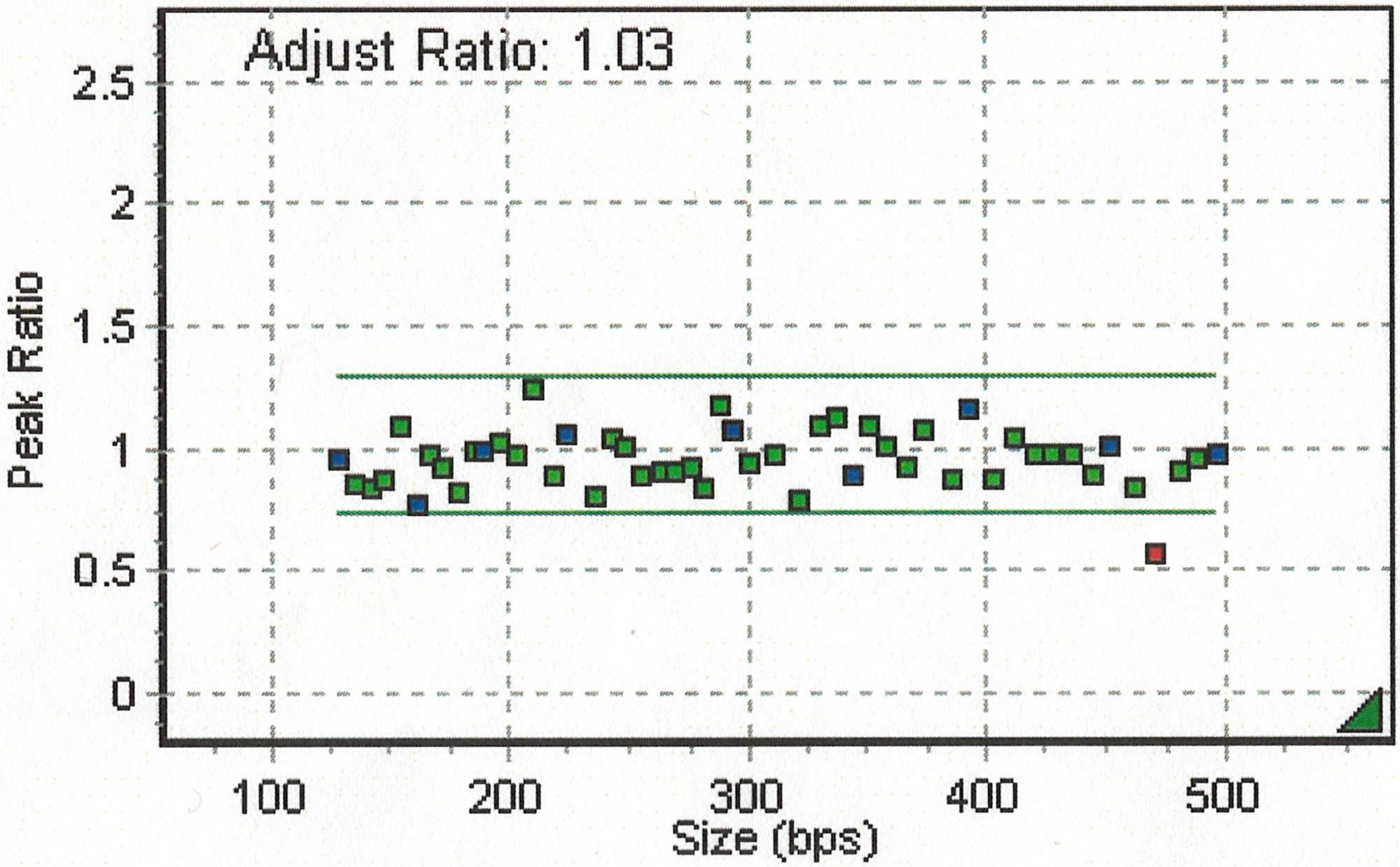


| Probe Name | Bin Size | MLPA |
|---|---|---|
| PALB2-E1 | 275.4 | 0.927 |
| PALB2-E2 | 248.9 | 1.007 |
| PALB2-E3 | 402.9 | 0.881 |
| PALB2-E4 | 287.5 | 1.188 |
| PALB2-E5 | 471.5 | 0.57 |
| PALB2-E6 | 373.6 | 1.083 |
| PALB2-E7 | 488.2 | 0.965 |
| PALB2-E8 | 420.3 | 0.972 |
| PALB2-E9 | 147.8 | 0.881 |
| PALB2-E10 | 172.3 | 0.927 |
| PALB2-E11 | 310.9 | 0.979 |
| PALB2-E12 | 462.1 | 0.845 |
| PALB2-E13 | 350.4 | 1.104 |
